# Assessing the lockdown effect from excess mortalities

**DOI:** 10.1101/2021.01.22.21250312

**Authors:** Alexej Weber

## Abstract

**Background and Aims:** The reported case numbers of COVID-19 are often used to estimate the growth rate of infections. We use the excess mortality instead to show the effect of most restrictive non-pharmaceutical interventions (mrNPIs) as compared to less restrictive NPIs (lrNPIs) concerning growth rate and death counts.

**Methods:** We estimate the COVID-19 growth rate for Austria, France, Germany, Italy, Netherlands, South Korea, Spain, Sweden, UK and USA from the excess mortality. We use the average growth rate obtained for Sweden and South Korea, the only countries with lrNPIs only, to estimate additional death numbers in the other countries, had mrNPIs not been applied.

**Results:** The growth rate estimated from excess mortality decreased faster for countries with mrNPIs than for Sweden and South Korea, suggesting that the mrNPIs do have a non-negligible effect. Implementing lrNPIs instead of mrNPIs results in up to 3 times higher death numbers. This is not visible when the growth rate is calculated using the reported case numbers of COVID-19 instead of the excess mortality.

**Conclusion:** The conclusion for the spreading of COVID-19 obtained from reported COVID-19 cases in previous studies are most likely biased. Using our method, a more realistic estimate of the growth rate is obtained. Conclusions made for the reproduction number derived from the reported case numbers, such as the apparent insignificance of mrNPIs (lockdowns), might therefore be wrong and will have to be reevaluated using the growth rates obtained with our method.

## Introduction

To face the spread of COVID-19 disease, many countries introduced the so-called non-pharmaceutical interventions (NPIs). The most restrictive NPIs (mrNPIs) policies include mandatory stay-at-home and business closures (lockdowns). Since the mrNPIs may have potential harmful side effects [1-13] the risk-benefit ratio has to be evaluated. Usually, either the COVID-19 deaths data or the result of positive testing is used to assess the spreading of the disease and calculate the effect of NPIs [14-16]. While [15-16] evaluate mrNPIs as crucial for saving millions of lives, [14] sees no significant effect of mrNPIs over lrNPIs. However, the data used in [14-16] is of poor quality for the following reasons:

- **First**, the testing may have started too late to trace the beginning of the epidemic, therefore early deaths and cases attributable to COVID-19 may have been missed.
- **Second**, the false positive rate and the country specific counting methods have to be taken into account. Usually, the testing amount increases over the period of several weeks. While the actual COVID-19 cases may fall on average, the number of positive tests will suggest the opposite situation.
- **Third**, even if the testing does not have any false positives and finds the actual SARS-COV2 virus fragments in individuals, it does not give us the information whether the virus amount found in individuals is sufficient to cause a severe disease. The very sensitive PCR-Tests might detect the same apparent spreading in different countries; however, in countries like Germany with mrNPIs, the virus amount in positively tested individuals might on average be smaller than in countries with lrNPIs. This would imply that in countries with mrNPIs, positive tested individuals are less infectious on average than in countries with lrNPIs. Looking solely at the positive test numbers, which may display a similar pattern in various countries, one then wrongly concludes that the spreading of the COVID-19 disease is equivalent.
- **Fourth**, also the testing results are subject to the local decision of who is to be tested and whether the deaths with a positive test outcome are to be counted as COVID-19 deaths or not, which may vary over time. If the testing were to be restricted to the risk group, reducing the number of total tests, the positive counts may drop suddenly. Changing the convention of deaths counting also leads to sudden structural breaks in the data.
- **Fifth**, also the testing is not standardized; so in the beginning, the positive tests may be repeated to exclude the false positives, while later on, when the laboratories work at their limits, the positive tests are not reevaluated, leading to a situation which appears to be much worse.

Although the reported deaths attributable to COVID-19 are more reliable data than data for positive test numbers, many of the problems mentioned above remain. Basically, it is not clear whether the person died from the disease or with the disease, leading to a biased estimate for the reproduction number or growth rate.

Due to the above challenges, we restrict our data to excess mortalities as this is the most solid number, being directly linkable to the spreading of the deadly disease.

The exact dynamics of the spreading of the SARS-COV2 virus in the population is unknown. In addition to bad initial data based on case numbers, the following effect may lead to over- or under-estimation of the effect of mrNPIs. During the first wave of the COVID-19 pandemic, many countries first implemented lrNPIs and then some countries implemented mrNPIs as well. Some assume that without mrNPIs the initial exponential phase would simply continue until everyone is infected. This however is not correct, as lrNPIs also have a substantial effect. Additionally, even without any NPIs, the virus spread will eventually go down due to changes in human behavior, so that the simple assumption of further exponential spreading without mrNPIs is not correct and will lead to an overestimation of death and infection numbers, as for example in [15-16]. Another drawback is the assumption that even without lockdowns the observed effect of lrNPIs would continue (which some researchers implicitly assume in their models, as in [14]). This assumption lets them argue that no additional benefit of mrNPIs is visible. However, it could be that the infection rate only behaved in the observed way precisely because of mrNPIs which were later installed.

Because of these problems, in this paper, we do not compare the spreading rate before and after the interventions in each country, but rather compare the shape of the excess mortality among different countries. The benefit of this approach is that we do not have to assume models for the effects of NPIs on infection rates, but rather calculate the growth rate directly from mortality data. We then can directly compare whether the growth rate in lrNPI countries behaves differently from mrNPI countries.

We perform the analysis for the same set of countries used in [14], with one exception. Since we do not have mortality data for Iran, we included Austria instead to have the same total number of countries. The countries analyzed in this work therefore are: Austria, France, Germany, Italy, Netherlands, South Korea, Spain, Sweden, UK and USA. South Korea and Sweden applied lrNPIs only. USA is a special case, as some states either used lrNPIs only or re-opened again soon after mrNPIs had been implemented. South Korea, in addition to lrNPIs, had advanced surveillance technology for tracking infections.

## Data

We used data from the human mortality data base [17] which gets the data from the regional statistical bureaus.

## Methods

To overcome the testing problems mentioned in the introduction, we propose to calculate the growth rate directly from the excess deaths obtained from weekly or daily mortality data. While this method will not work when the excess deaths are small compared to the average noise level, it provides a reasonable result when the excess deaths are much bigger than noise and can be attributed to the spreading of the disease, causing the individual to die within a few weeks after infection. This is the case for COVID-19 in the first half of the year 2020, where we do not expect to see big side effects from NPIs yet (compared to COVID-19 itself), but where COVID-19 in fact lead to a big excess mortality. Although the method can also be applied to the second half of the year, additional side effects of NPIs have to be carefully estimated, and cannot be neglected.

To calculate the excess deaths, the actual deaths have to be subtracted from the expected deaths. Usually, the expected deaths are calculated by taking the average of several past years of data, or by applying a more sophisticated statistical model to past years [18-22]. Since the weekly death data is available for a few past years only, the expected deaths are usually calculated by taking around 4-10 past years into account. This direct approach leads to an overestimation of the expected deaths, independently of the statistical method used, due to influenza and heat waves. Since the influenza waves mostly occur around the beginning of the year, they do not cancel out. The statistical model interprets the influenza wave as a regular pattern and the estimate of the expected deaths is biased. Instead, the expected deaths have to be estimated from clean years without a strong influenza wave adjusted to the current level of deaths, accounting for a crisis-free baseline.

Like influenza, the COVID-19 disease is more deadly to the elderly population and to people with serious preconditions. Deaths of these people form part of the expected deaths curve. If they are hit by the disease and die, the expected deaths will be lowered for some period after the disease wave, since the most vulnerable have “pre-died” some weeks earlier (“harvesting effect”), see [23, 24].

Additionally to directly deriving the growth rate from excess mortality numbers, we also considered the impact of the harvesting (pre-dying) effect by distributing a portion of the cumulated excess deaths equally to a period of 20 weeks into the future, thus lowering the baseline of the year, see [24] for more details. In this way, we obtain more excess deaths compared to the case when the actuals deaths are subtracted from the baseline year directly. We have chosen 20 weeks, since we observe the lowering of deaths numbers after the COVID-19 wave for a period of about 10 weeks, and the first COVID-19 wave lasts for around 10 weeks itself.

We observe a potential harvesting effect in all countries which implemented mrNPIs (except for USA). This means that the excess mortality during the first COVID-19 wave was followed by a mortality deficit. In countries which implemented lrNPIs (and in the USA), we do not observe the mortality deficit after the COVID-19 wave. This could be due to two main reasons: Either mostly healthy people died (so no pre-dying effect), or the COVID-19 wave is still continuing in the period after the excess mortality, so that although the death numbers correspond to the expected value, still a fraction is dying from COVID-19. We favor the second interpretation as the first one is highly unlikely – the virus should hit on average the same population groups equally.

We therefore calculate the growth rate with and without a harvesting effect.

To calculate the baseline year, we did the following: We chose clean years, adjusted them to the starting level of 2020, and applied a lower order moving average to reduce the remaining noise.

For most countries, we constructed the base line from the years 2016 and 2019 as other years contained strong influenza waves. For South Korea, we took 2017 and 2019.

To calculate the growth rate, we take the first difference (corresponding to first derivative) of the logarithm [25-26] of the excess deaths (which we assume to be due to COVID-19). We also lag the result with 2 weeks, to obtain the corresponding growth of infections (assuming that it takes 1 week after the infection to develop a disease and 1 week for the individual to die from the disease). Since the data is noisy, we finally also smooth the excess mortality and the growth rate number using a lower order moving average.

## Results

The mortality deficit in all mrNPI countries right after the first COVID-19 wave can be well explained with the harvesting effect, where 30-70% pre-die in the wave, namely those, mostly elderly, individuals with short life expectancy of around 20 weeks (who would have died otherwise in the mortality deficit period). We thus calculate the results for the representative values of 0 (no pre-dying at all) and 0.5 (50% of counted deaths in the excess period are attributed to pre-dying). The pre-dying leads to a broader and bigger excess mortality since the baseline mortality is lowered (see [24]). Thus, the calculated infection growth rates decay slower if the pre-dying effect is taken into account. However, the qualitative results of this study are unchanged by the pre-dying effect.

Figure 1 demonstrates the lowering of the base line for Germany with a pre-dying effect of 70%, where it explains the mortality deficit in calendar weeks 19 – 29.

**Figure 1.**
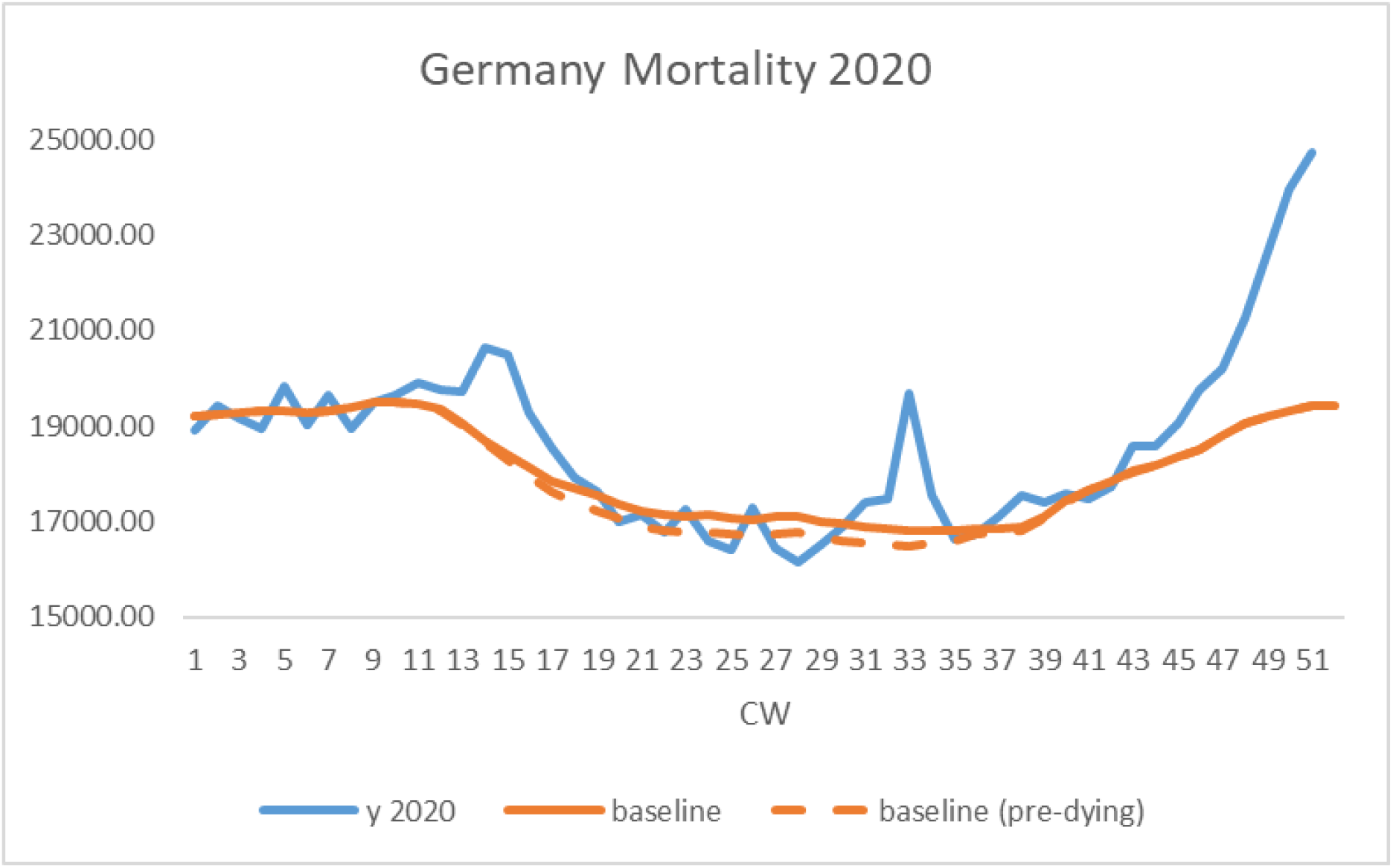
The mortality curve is shown for Germany (blue). The prominent COVID-19 wave in weeks 10 – 19 is followed by a mortality deficit in weeks 19-29, where the mortality data on average is lower than the baseline mortality (solid orange line). The mortality deficit gets interrupted by the heat wave in weeks 30 – 35. Then in week 40, the second COVID-19 wave starts. Assuming that during the first COVID-19 wave around 70% of deaths can be attributed to pre-dying, we can explain the mortality deficit in weeks 19-29 by lowering the baseline, i.e. a part of individuals with expected death date in the near future pre-dies in the COVID-19 wave and leaves the base line curve.

When the growth rates are estimated from the positive test cases as in [14], it seems that the spreading never stops and after the initial exponential phase continues in a linear fashion at a constant but very high rate, although the death numbers fall dramatically.

In contrast, when the growth rates are obtained from the excess mortalities as in this study, we see that the spreading was exponential in the beginning and then dropped and decreased exponentially. The representative results of the behavior of the growth rate (for Germany, Italy, Sweden and South Korea) are shown in Figure 2. For this figure, we have chosen two representative countries with mrNPIs (Germany hit much less than Italy) and two countries with lrNPIs (South Korea hit much less than Sweden).

**Figure 2.**
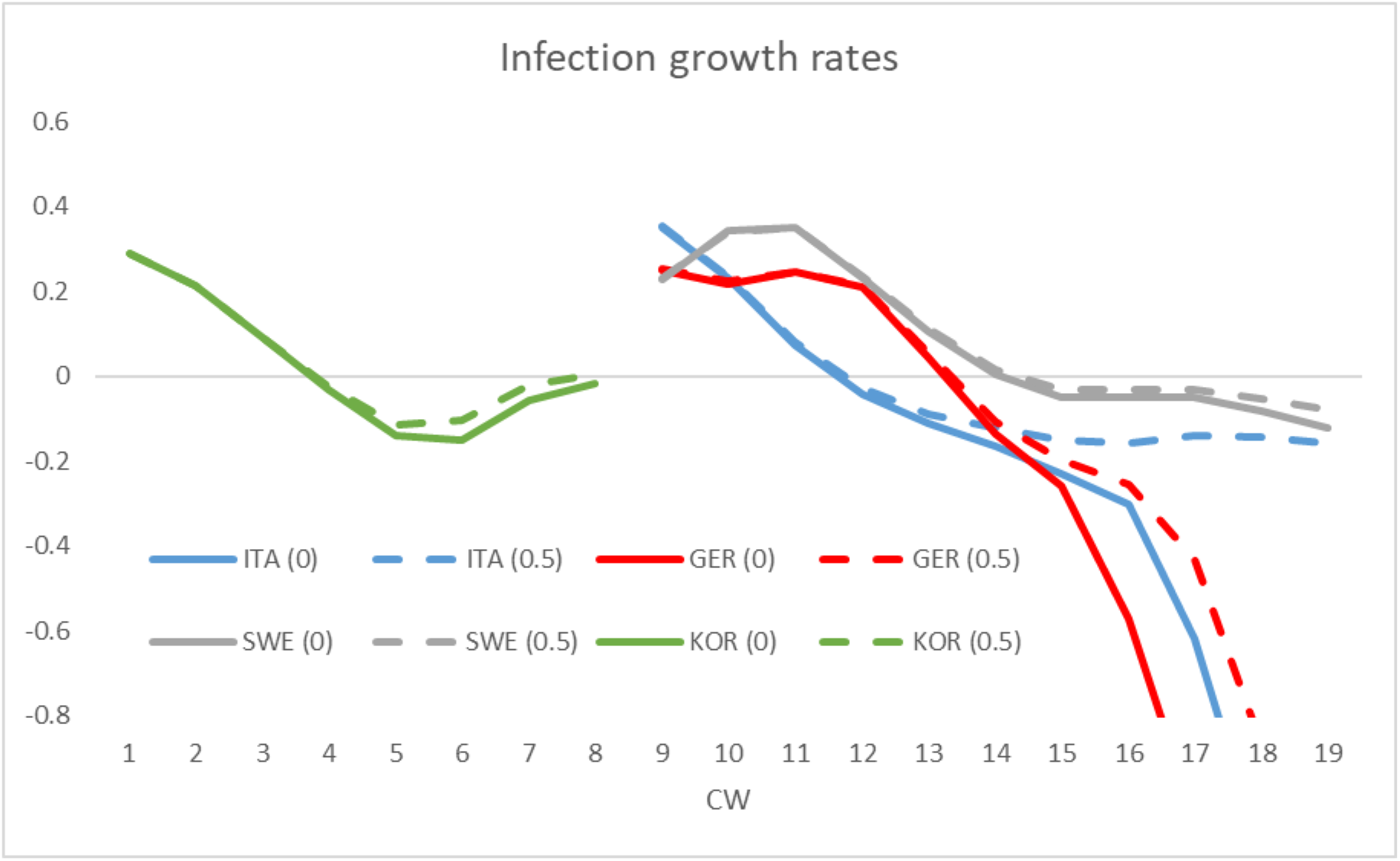
The figure shows the infection growth rates as deduced from excess mortalities, for Italy, Germany, Sweden and South Korea for two parameter values of pre-dying 0% (solid lines) and 50% (dashed lines). The countries with mrNPIs (Germany and Italy) have lower growth rates on average, once the growth rate is below 0 (corresponding to mrNPIs becoming implemented). All curves start at a high positive value, corresponding to the high exponential spreading in the beginning of the disease wave.

In Figure 2, we see, that Italy and Germany, which installed mrNPIs, experienced a more negative growth rate than South Korea and Sweden with only lrNPIs. This means, that the new infections decayed much faster in Germany and Italy right after the mrNPIS had been installed (Italy implemented early lockdowns from week 9 and a national quarantine from week 11, Germany from week 13). In contrast, in Sweden and South Korea (without mrNPIs), the drop from 0 soon stops at a higher value. This means that the excess mortality curve is wider due to a much weaker exponential decay.

For the countries analyzed in this study, mrNPIs were always implemented when the growth rate was in the positive range, right before the zero value.

In order to investigate, whether the growth rate would decline faster in countries with mrNPIs as compared to countries with lrNPIs, we calculated the average growth rate after the growth rate had passed the zero value (for the following 4 weeks on average) for all countries (with and without mrNPIs). More negative values correspond to faster decrease of new infections and new death numbers. We chose a period of 4 weeks, as during this period all countries display sufficiently large excess mortality. The results are summarized in Table 1.

**Table 1.**
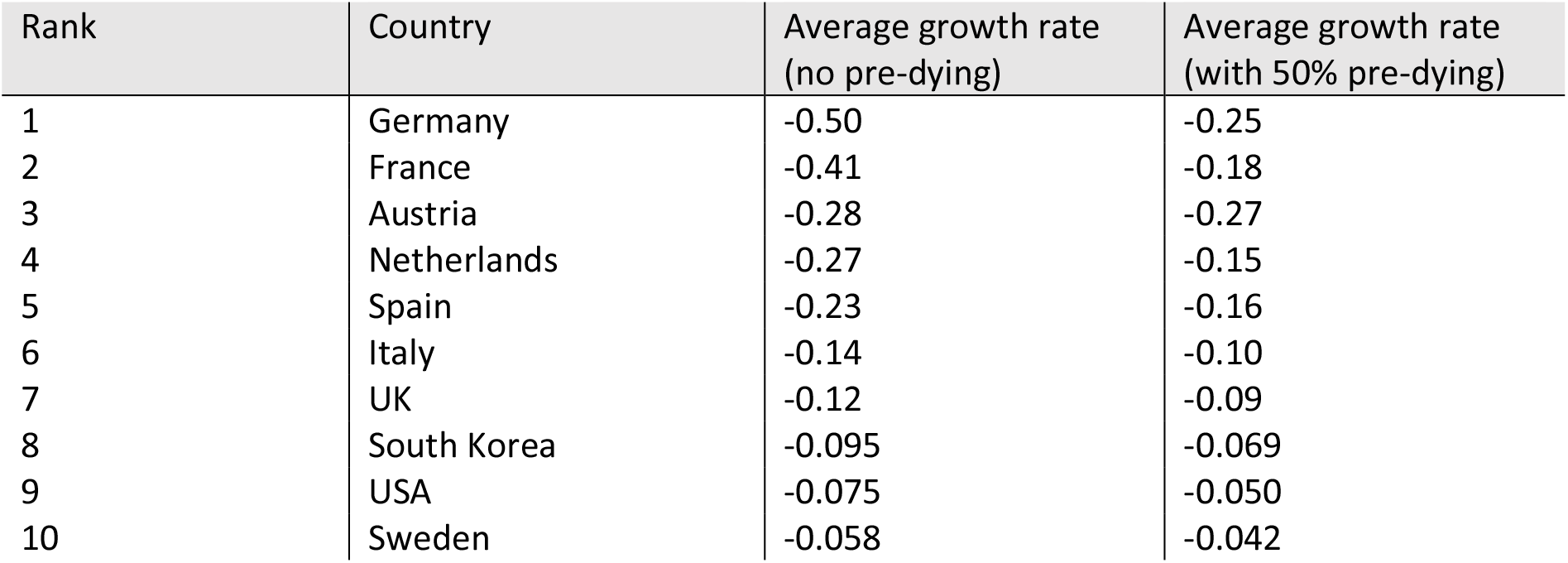
The table shows the growth rate in the decaying period averaged over 4 weeks. The countries are ranked according to the growth rate with no pre-dying effect. The countries with lrNPI (South Korea and Sweden) and USA occupy last places.

| Rank | Country | Average growth rate<br>(no pre-dying) | Average growth rate<br>(with 50% pre-dying) |
| --- | --- | --- | --- |
| 1 | Germany | -0.50 | -0.25 |
| 2 | France | -0.41 | -0.18 |
| 3 | Austria | -0.28 | -0.27 |
| 4 | Netherlands | -0.27 | -0.15 |
| 5 | Spain | -0.23 | -0.16 |
| 6 | Italy | -0.14 | -0.10 |
| 7 | UK | -0.12 | -0.09 |
| 8 | South Korea | -0.095 | -0.069 |
| 9 | USA | -0.075 | -0.050 |
| 10 | Sweden | -0.058 | -0.042 |

Table 1 lists the average growth rate (averaged over 4 weeks) in the negative regime right after crossing the zero value assuming no pre-dying effect or a pre-dying of 50%, respectively.

We clearly observe that countries with mrNPI are ranked higher and countries with lrNPI occupy last positions. One exception is USA on the second last place. In this country, we find a mixed strategy: not every state implemented mrNPIs. Some states with mrNPIs also did open up again early. On the whole, USA has had many different local interventions with no clear global strategy. Korea is ranked higher than USA and Sweden, the growth rate decayed faster probably due to its advanced infection tracking technology. Sweden with a clear lrNPI strategy occupies the last position.

It can furthermore be observed, that once the pre-dying is taken into account, the ranking slightly changes among mrNPI countries (especially in the top ranks), but this does not change the qualitative picture that the countries with lrNPIs are ranked last. In fact, if we average the results of the calculation with or without pre-dying, the ranking is equivalent to that in Table 1.

On average, mrNPIs had around three times as low spreading rates as compared to the lrNPIs countries. The mean value of the growth rate for countries with mrNPIs is -0.25 [-0.37,-0.13] for 0% pre-dying. Taking a 50% pre-dying effect into account, the mean value reduces to -0.16 [-0.22,-0.09]. In contrast, the mean value for South Korea and Sweden is -0.076 for 0% pre-dying and -0.054 for 50% pre-dying. We calculated the confidence intervals by taking country averages.

Although sample size is small, (we only have data for 2 countries with lrNPIs, and thus are not able to calculate the confidence bands for the lrNPI mean value if working with country averages), the very low ranking of all study countries with lrNPIs still suggests that mrNPIs had a non-negligible effect on the spreading of COVID-19. Working with 4 growth rate values per country, however, we are able to calculate the confidence intervals for mrNPI and lrNPI mean values. These are the results obtained by the second method: -0.25 [-0.34,-0.16] (mrNPI, for 0% pre-dying) and -0.16 [-0.20,-0.11] (mrNPI, for 50% pre-dying), -0.076 [-0.114, -0.039] (lrNPI for 0% pre-dying) and -0.054 [-0.085, -0.022] (lrNPI, for 50% pre-dying). This suggests a significant effect of mrNPIs over lrNPIs as the corresponding confidence intervals do not overlap.

To find out what this result means for the death numbers in countries with mrNPIs, we replace the negative growth rates for countries with mrNPIs with the corresponding average numbers obtained for Sweden and South Korea, thus simulating a lrNPI scenario for a period of 6 weeks. We then recalculate the resulting additional death numbers and the corresponding percentage increase with respect to original excess mortality in that period. The results are listed in Table 2. We see that the percentage increase of death numbers differs vastly between countries, being biggest (around 170%) for Germany (where the decay of the infection growth rate was biggest under mrNPI) and smallest (3%) for USA (where the decay of the infection growth rate was smallest under mrNPI).

**Table 2.**
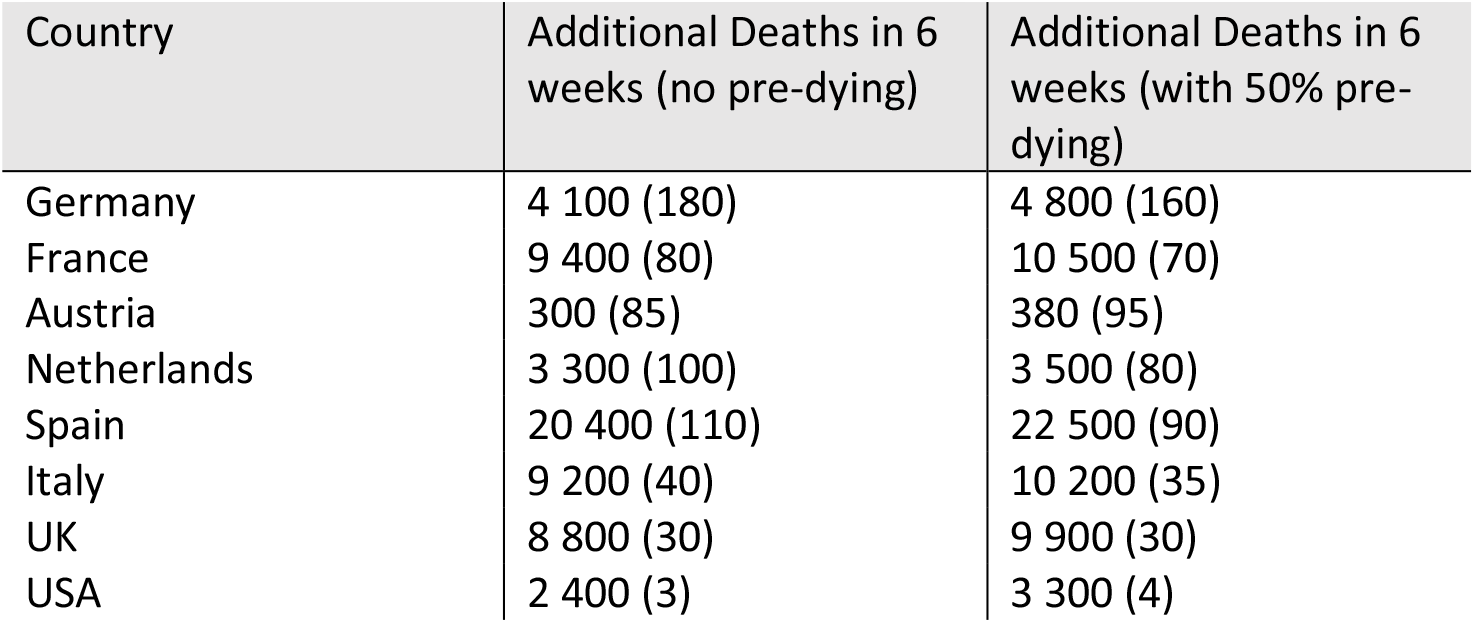
The table shows the additional death numbers for a period of 6 weeks (together with the corresponding percentage increase with respect to original excess mortality in that period), assuming the countries had not implemented mrNPIS but kept lrNPIs instead. Values taking the pre-dying into account are given in the third column. (The apparent sudden percentage jump from Germany to France and from Spain to Italy is due to discrete nature of weekly data used)

The effect on the excess mortality if lrNPIs had been in place instead of mrNPIs is shown Figure 3 for Italy and Germany as representative examples. We see that due to a substituted less negative growth rate, the decay of new infections and thus the new death numbers is slowed down. The prolongation of the excess mortality leads to additional deaths (for the 50% pre-dying scenario, the effect is qualitatively the same, so we do not show it).

**Figure 3.**
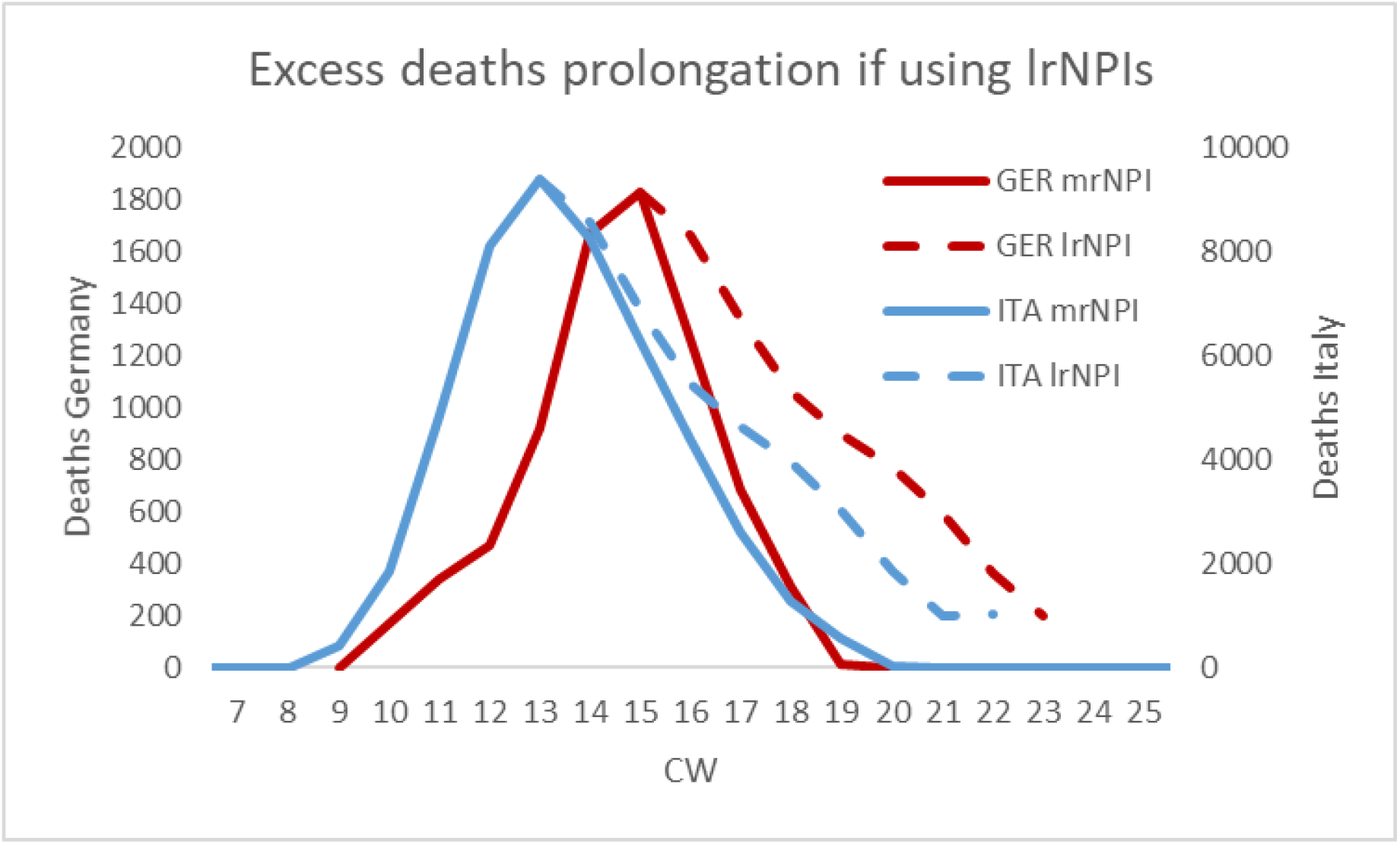
The figure shows the original excess mortality for Germany and Italy (solid lines, left and right y-Axis, respectively) for 0% pre-dying value. Had lrNPIs been used instead (scenario dashed line), the growth rates would be higher and the excess mortality curve accordingly prolonged in the corresponding country. The wider excess mortality curves would have resulted in ca. 4 000 more deaths for Germany and ca. 9 000 more deaths for Italy during the 6 weeks under the lrNPIs scenario.

## Conclusion

We demonstrated that when calculating the growth rate from the excess mortality instead of the reported case numbers, the mrNPI effect is clearly visible. Countries which implemented mrNPIs were capable of stopping the spread of SARS-COV2 in the population more effectively than Sweden and South Korea on average by decreasing the spread exponentially. With lrNPIs, the exponential decrease was not very pronounced compared to countries having mrNPIs. The reason the effect was not visible in previous studies such as [14] may have two main sources. First, misleading data: extended testing shows increasing numbers of people to be infected, while the average prevalence may have dropped. Second, wrong models, which implicitly assumed that even without lockdowns, the observed effect of lrNPIs would continue.

Both sources mask the effect of the mrNPIs over lrNPIs.

## Data Availability

all data is publically available under the following links listed below

https://www.mortality.org

## Notes

### Competing Interest Statement

The authors have declared no competing interest.

### Clinical Trial

not relevant, since not a clinical study

### Funding Statement

NA since no funding

### Author Declarations

NA since independent researcher

### Summary of Updates

Extented statistical analysis (more study countries), updated data, additional tables, figures and text for more clarity,

